# The Challenges for Screening and Diagnosing Gestational Diabetes Mellitus in Brazil: a cross-sectional study in a low-risk obstetric hospital

**DOI:** 10.1101/2022.01.08.22268941

**Authors:** Bruna Marcolino Paes Maria, Debora Giovanna Fernandes Vivaldo, Thais Sales Izidoro, Ana Paula Costa De Freitas, Luiz Takano, Talita Domingues Caldeirao, Carlos Izaias Sartorao Filho

## Abstract

**Background:** Gestational Diabetes Mellitus (GDM) is a very prevalent disease and can cause several morbidities for women and their offspring. The literature demonstrates the necessity for a better approach during prenatal assistance to detect and treat the disease. We aimed to evaluate the model and efficacy of GDM screening and diagnosis in a referenced low-risk obstetrical center of the municipality of Assis, Sao Paulo state, Brazil. Moreover, the specific objective was to evaluate the prevalence of GDM.

**Methods:** We conducted a retrospective cross-sectional study of pregnant women, in which 257 prenatal cards and the clinical approaches used for GDM diagnosis and their results. We observed the consecutive patients admitted to the low-risk referenced obstetrical service of the “Santa Casa de Assis-SP” for childbirth from January to August 2021.

**Results:** There were 257 pregnant women, 227 prenatal cards obtained. Of these, 24.6% of the cards were considered incomplete, 25 (9.72%) did not contain the initial fasting plasma glucose information, and 93 (36.18%) did not describe this information in the second to the third trimester. The prevalence of GDM in the population was 14.78%.

**Conclusion:** We observed many pregnant women not screened according to the recommended guidelines and many prenatal cards with incomplete information. According to the screening and diagnosis guidelines, GDM prevalence was underestimated. The lack of prenatal card information and inadequacy of screening and diagnoses were observed in this population.

## INTRODUCTION

Gestational diabetes mellitus (GDM) is defined as glucose intolerance of variable degree with onset or first recognition during pregnancy. (1) Epidemiological data indicate that GDM is one of the major metabolic disorders of pregnancy and that it has an estimated prevalence between 3 and 25%, varying according to the population studied and the diagnostic criteria used. (2) In the Brazilian Unified Health System (SUS), it is estimated that its prevalence range from 10 to 25%. Screening for GDM is universal and is part of the Prenatal Assistance Program of the Unified Health System, which is based on a set of measures and protocols of conduct, aiming at the early detection and intervention of factors that can lead to maternal-fetal complications. The guidelines proposed for the Brazilian Unified Health System are the same proposed by the World Health Organization (WHO).(4) However, there is an obstacle between the use of health services provided by pregnant women, difficulties in carrying out family planning, inadequate appointment scheduling, and instructions, lack of physical, emotional, and social support for these users by the multidisciplinary team. The obstacle in the standardization of the GDM approach becomes clear. (5) That said, as GDM is the most prevalent endocrinopathy of pregnancy-associated with its serious maternal-fetal complications, (6) it is essential to evaluate and standardize the screening and diagnosis proposed by the Brazilian SUS for GDM in the municipality of Assis-SP. Moreover, the approach to GDM is frequently recognized as inconsistent. (7) Despite a great amount of knowledge, the most appropriate diagnostic strategy, screening policy, and treatment options for pregnancies complicated with GDM remain uncertain in the medical community. (8)

We aimed to appraise the current regional practices of screening, diagnosis GDM in a referenced low-risk obstetrical center of the municipality of Assis, Sao Paulo state, Brazil. Moreover, the specific objective was to evaluate the incidence of GDM in pregnant women of this institution.

## METHODS

We performed a cross-sectional study on September 2021 from a retrospective secondary data source based on the Prenatal cards, and prenatal exam results of pregnant women admitted for the delivery at Santa Casa de Assis-SP. A retrospective survey of the hospital files collected sociodemographic information, and the model of screening or approach for GDM of each pregnant woman admitted from January to August 2021, namely: fasting glucose values in the first and second to the third trimesters and 75grams Oral Tolerance Glucose Test (OGTT), number of prenatal consultations, in addition to the quality and quantities of the described information given in the prenatal card.

The study was carried out at Santa Casa in Assis-SP due to the obstetrical low-risk reference hospital in the city for the performance of obstetric childbirths through the Brazilian Unified Health System (SUS). The Study was approved by the Ethics Committee of Educational Foundation of Assis Municipality under number CAAE: 42185120.4.0000.8547.

The analyzed group consisted of pregnant women over 18 years old admitted to the Santa Casa de Assis, for childbirth, through the SUS.

We included: Pregnant women over 18 years old, admitted for childbirth through SUS, from January to August 2021, at Santa Casa de Assis-SP. We excluded: Pregnant women with type 1 or 2 diabetes mellitus diagnosed before the pregnancy, twin or more pregnancies, previous history of bariatric surgery, history of hypoglycemic drugs before the pregnancy.

Data were collected anonymously, and its disclosure took place in a compiled manner to minimize the risk of breaching anonymity and confidentiality.

Evaluation of the screening and diagnosis of GDM and identifying weaknesses in the information provided by the prenatal cards can provide educational actions with health teams to standardize and improve the accuracy of the diagnosis of this important disease.

Sociodemographic data analyzes the population’s rate that performs the screening for GDM as recommended, and the prevalence of GDM was estimated according to the criteria currently recommended by the Brazilian Guidelines for Screening and Diagnosis of GDM. The data obtained were compiled and arranged in spreadsheets for statistical analysis.

The data collected from the pregnant woman’s cards and the medical records provided by Santa Casa de Assis were obtained from the obstetrical recorded files of the institution. The variables of interest were: maternal age in years, number of prenatal consultations recorded in the card, gestational age at the admission for childbirth in weeks, maternal weight at the first prenatal visit and the birth in Kilograms and the Body Mass Index, ethnicity (white, black, brown or Asiatic), Married/partner relationship and not Married or no partner relationship, Religion (catholic, protestant, and others), the adequacy of prenatal card records (yes or not, according to legibility and quantity and quality of information described). In addition, fasting plasma glucose in the first trimester (yes or not, and the value in mg/dL), fasting plasma glucose in the second/third trimester (yes or not, and the value in mg/dL), 75 grams OGTT (yes or not described and the values in mg/dL of the fasting, 1 hour and 2 hours test), the described diagnosing of GDM in the current pregnancy (yes or not). In addition, we considered the WHO / Brazilian Health Ministry guidelines for GDM screening and diagnosis.

We performed a descriptive data analysis of the variables using the IBM-SPSS software, version 25.

## RESULTS

A total population of 271 participants was recovered, surpassing the sample size calculation of 196 participants for a 5% margin of error and a 95% confidence interval. The population clinical and demographic characteristics data are provided in Tables 1 and 2. A total of 257 eligible pregnant women were analyzed, of which 227 presented their prenatal care cards. Of these, 24.6% of the cards were considered incomplete, 25 (9.72%) did not contain the fasting plasma glucose information from the first trimester, and 93 (36.18%) did not describe this information in the second/third trimester. Of 233 first-trimester fasting blood glucose results, 29 (12.44%) were positive for GDM. Fourteen patients were diagnosed in the second trimester using the fasting plasma glucose value equal to or more than 92 mg/dL or the 75-g OGTT positive for GDM. Considering 60 patients who underwent 75g OGTT, GDM was diagnosed in 12 patients (20%) according to the WHO criteria, 7 of which were diagnosed at the basal fasting plasma glucose, 3 in the first-hour test, and 2 in the second-hour test, after ingestion of 75g of dextrose. In 167 cards, there was no information about the screening with the recommended 75g OGTT test. The GDM diagnosis, according to the Fasting Plasma Glucose equal or more than 92 mg/dL in the first trimester and then, if this test was negative, the 75g OGTT positive in the second to the third trimester, totalized 38 of 257 participants (14.78%).

**Table 1.** Quantitative Demographical and clinical characteristics.

|  | N | % |
| --- | --- | --- |
| **White** | 171 (257) | 66,67 |
| **Brown, Black or Asian** | 86 (257) | 33.33 |
| **Married or living with a partner** | 194 (257) | 75.18 |
| **Not married / no partner** | 63 (257) | 24.82 |
| **Catholic** | 113 (257) | 44.33 |
| **Protestants** | 91 (257) | 35.46 |
| **Other religions** | 53 (257) | 20.21 |
| **Prenatal card** | 227 (257) | 88.30 |
| **Adequated Prenatal card** | 171 (227) | 75.33 |
| **Inadequated Prenatal card** | 56 (227) | 24.66 |
| **1st trimester hyperglycemia** | 29 (233) | 12.44 |
| **FPG 1st trimester not informed or absent** | 25 (257) | 9.72 |
| **Performed 75g OGTT** | 60 (257) | 23.34 |
| **2nd trimester not informed or absent** | 93 (257) | 36.18 |
| **2nd trimester FPG positive** | 13 (233) | 5.57 |
| **1 hour 75g OGTT positive** | 3 (60) | 1.16 |
| **2- hour 75g OGTT positive** | 2 (60) | 3.33 |
| **GDM \*** | 38 (257) | 14.78 |

FPG: Fasting Plasma Glucose
75g OGTT: 75 grams Oral Glucose Tolerance Test
\*WHO criteria

**Table 2:** Qualitative Demographical and Clinical characteristics. N = 257

|  | Mean | Standard deviation | Minimum | Maximum |
| --- | --- | --- | --- | --- |
| **Age (years)** | 26.23 | 5.54 | 18.00 | 42.00 |
| **Number of Prenatal visits** | 8.73 | 2.68 | 1.00 | 16.00 |
| **Gestational Age (weeks)** | 38.54 | 1.71 | 27.00 | 42.00 |
| **Prepregnancy Maternal Weight (Kg)** | 70.19 | 15.66 | 39.00 | 130.00 |
| **Maternal Weight at birth (Kg)** | 79.99 | 14.07 | 46.00 | 127.80 |
| **Maternal weight gain (Kg)** | 9.8 | - 1.5 | 7.00 | - 2.2 |
| **Height (m)** | 1.62 | 0.06 | 1.34 | 1.76 |
| **BMI prepregnancy (Kg/m<sup>2</sup>)** | 26.68 | 5.56 | 15.82 | 50.94 |
| **BMI at birth (Kg/m<sup>2</sup>)** | 30.34 | 5.16 | 18.73 | 50.55 |
| **BMI gain (score)** | 3.66 | 5,16 | -3.90 | 2.91 |
| **FPG 1st trimester (mg/dL)** | 81.71 | 8.20 | 60.00 | 107.00 |
| **FPG 2nd trimester (mg/dL)** | 78.03 | 11.53 | 56.00 | 144.00 |
| **FPG 75g OGTT (mg/dL)** | 80.43 | 17.44 | 52.00 | 134.00 |
| **1-hour Plasma Glucose (mg/dL)** | 109.35 | 42.25 | 60.00 | 194.00 |
| **2-hours Plasma Glucose (mg/dL)** | 89.02 | 43.97 | 56.00 | 164.00 |

FPG: Fasting Plasma Glucose
75g OGTT: 75 grams Oral Glucose Tolerance Test

## DISCUSSION

We observed many pregnant women not screened according to the recommended guidelines and many prenatal cards with incomplete information. As a result, according to the screening and diagnosis guidelines, GDM prevalence was underestimated. Furthermore, the information obtained from the prenatal cards of pregnant women attended to childbirth at the Santa Casa de Assis-SP described a lack of uniformization in the GDM approach in the municipality, although the number of prenatal visits was adequate. As a result, a prevalence of GDM of 14.78%. In a recent meta-analysis involving a great number of pregnant women (136705) in 31 different studies, the GDM prevalence using the current screening and diagnosis method was 14.7%, a 75% of increment when compared with the oldest screening methods. (9)

Because of the significant number of patients affected by GDM, even in a population with a considered number of non-adequately screening, we presume that pregnant women’s early and universal screening is essential in performing early diagnosis and intervention and thus avoiding future maternal-fetal complications. Besides, we presume that the prevalence of the GDM is higher than observed, especially considering the higher number of overweight and obesity. Furthermore, we observed many pregnant women not screened according to the recommended guidelines and many prenatal cards with incomplete information.

Concerning limitations, the associations identified were difficult to interpret. More, the variables were susceptible to bias due to inadequate response and misclassification. In addition, we did not focus on the maternal, fetal, and neonatal GDM associated outcomes, which could be essential to evaluate our population.

Although our findings are parochial, their implications may be global. Other studies worldwide demonstrate the professional confusion and difficulties due to the different GDM conception, classification, and models of screening and diagnosing. (7,10,11)

## CONCLUSION

We demonstrated the parochial lack of opportunity for GDM screening and diagnosis in a city of Brazil, despite the current guidelines regarding the standardization for the disease. Different efforts to improve the knowledge and perceptions of the caregivers and further medical education to standardize the protocols would make the discordant approach to GDM more harmonious and improve the obstetric care of pregnant women.

Our findings are crucial to alert health system managers to propose strategies to improve the quality of screening and diagnosis for GDM.

## Data Availability

All data produced in the present work are contained in the manuscript

